# Automatic segmentation of spinal cord lesions in MS: A robust tool for axial T2-weighted MRI scans

**DOI:** 10.1101/2025.01.22.25320959

**Authors:** Enamundram Naga Karthik, Julian McGinnis, Ricarda Wurm, Sebastian Ruehling, Robert Graf, Jan Valosek, Pierre-Louis Benveniste, Markus Lauerer, Jason Talbott, Rohit Bakshi, Shahamat Tauhid, Timothy Shepherd, Achim Berthele, Claus Zimmer, Bernhard Hemmer, Daniel Rueckert, Benedikt Wiestler, Jan S. Kirschke, Julien Cohen-Adad, Mark Mühlau

## Abstract

Deep learning models have achieved remarkable success in segmenting brain white matter lesions in multiple sclerosis (MS), becoming integral to both research and clinical workflows. While brain lesions have gained significant attention in MS research, the involvement of spinal cord lesions in MS is relatively understudied. This is largely owed to the variability in spinal cord magnetic resonance imaging (MRI) acquisition protocols, high individual anatomical differences, the complex morphology and size of spinal cord lesions - and lastly, the scarcity of labeled datasets required to develop robust segmentation tools. As a result, automatic segmentation of spinal cord MS lesions remains a significant challenge. Although some segmentation tools exist for spinal cord lesions, most have been developed using sagittal T2-weighted (T2w) sequences primarily focusing on cervical spines. With the growing importance of spinal cord imaging in MS, axial T2w scans are becoming increasingly relevant due to their superior sensitivity in detecting lesions compared to sagittal acquisition protocols. However, most existing segmentation methods struggle to effectively generalize to axial sequences due to differences in image characteristics caused by the highly anisotropic spinal cord scans. To address these challenges, we developed a robust, open-source lesion segmentation tool tailored specifically for axial T2w scans covering the whole spinal cord. We investigated key factors influencing lesion segmentation, including the impact of stitching together individually acquired spinal regions, straightening the spinal cord, and comparing the effectiveness of 2D and 3D convolutional neural networks (CNNs). Drawing on these insights, we trained a multi-center model using an extensive dataset of 582 MS patients, resulting in a dataset comprising an entirety of 2,167 scans. We empirically evaluated the model’s segmentation performance across various spinal segments for lesions with varying sizes. Our model significantly outperforms the current state-of-the-art methods, providing consistent segmentation across cervical, thoracic and lumbar regions. To support the broader research community, we integrate our model into the widely-used Spinal Cord Toolbox (v7.0 and above), making it accessible via the command sct deepseg `-task seg sc ms lesion axial t2w -i <path-to-image.nii.gz>`.

## 1 Introduction

Multiple sclerosis (MS) is a complex autoimmune disease affecting the central nervous system, including the brain and spinal cord (Jakimovski et al., 2024). With over 2.8 million people impacted worldwide, MS is a leading cause of severe neurological disability in young adults (Walton et al., 2020). The disease is characterized by inflammatory demyelinating lesions, which are visible as hyperintensities on T2-weighted (T2w) MRI scans. Consequently, MRI has become the primary tool for diagnosing (Thompson et al., 2018) and monitoring MS (Rocca et al., 2024). However, most MRI studies focus on the brain and the measurement of atrophy in its anatomical regions (Calabrese et al., 2010; Kidd et al., 1999; Lucchinetti et al., 2011), despite spinal cord lesions being integral to MS diagnosis (Kearney et al., 2015; Thompson et al., 2018) and contributing significantly to disability (Bussas et al., 2022; Lauerer et al., 2024; Rocca et al., 2023).

Spinal cord MRI poses unique challenges due to its small size, tubular structure, proximity to moving organs, and the heterogeneous composition of surrounding tissues (Jasperse, 2024). These factors make lesion detection and manual segmentation cumbersome, time-consuming, and prone to variability (Gros et al., 2019). Existing open-source segmentation methods (De Leener et al., 2014, 2015; Gros et al., 2019; Naga Karthik et al., 2024) trained on specific regions of the spine fail to generalize under shifts in the data distribution caused by highly anisotropic axial scans. Furthermore, methods developed for segmenting brain lesions (Gentile et al., 2023; Mendelsohn et al., 2023; Schmidt et al., 2012; Shiee et al., 2010; Wiltgen et al., 2024) do not necessarily generalize to those of the spinal cord. In this study, we adopt an approach that simultaneously segments the spinal cord and lesions, focusing on axial T2w scans. Axial scans are superior for lesion detectability and can enhance diagnostic certainty in differentiating MS from other diseases, especially when high sensitivity for spinal cord lesions is required to confirm findings from sagittal scans or to detect small lesions potentially missed on sagittal sequences (Breckwoldt et al., 2017; Galler et al., 2016; Weier et al., 2012). Such protocols are increasingly applied in clinical practice (Kearney et al., 2015), although they require extended scanning times and, usually, acquisition in three chunks covering the cervical, upper thoracic, and thoracolumbar part of the spine; for brevity, we will refer to the latter two simply as thoracic and lumbar (part of the spine).

Automated spinal cord lesion segmentation presents several additional challenges. The appearance of lesions along with their ambiguous boundaries due to partial volume effects in anisotropic scans poses a significant challenge to the generalizability of segmentation models. Still, available training data (i.e., images with manually annotated reference standard lesion masks) are sparse, with a particular shortage for the thoraco-lumbar spinal cord. This is critical because, although most lesions develop in the cervical cord, a considerable proportion occurs in the thoracic spinal cord (Eden et al., 2019; Hua et al., 2015; Poulsen et al., 2021). Even in the most caudal part of the spinal cord (i.e., in the medullary conus), lesions can occur and be decisive for the individual patient regarding disability and differential diagnosis (Dubey et al., 2019; Mariano et al., 2021). In addition, it is unclear which data preprocessing^1^ and training strategies lead to the most optimal delineation of lesions. For instance, given the similar lesion intensities within axial slices across vertebral levels, does segmentation performance increase through additional training data from *other (possibly even distant) spinal levels*? In addition to the standard procedure of training on individual chunks, this can be explored by obtaining (stitched) whole spinal cord datasets and training a 3D model, which uses more contextual information surrounding the spinal cord (compared to a 2D model). Further, *how does the curvature of the spinal cord impact the segmentation performance*? Can *straightening*, a preprocessing strategy that virtually erects the spinal cord to a vertical column, removing curvature-related variance, (De Leener, Mangeat, et al., 2017) simplify the lesion segmentation task?

Lastly, given previous studies have primarily focused on the automatic segmentation of MS lesions in cervical or cervico-thoracic spine (Eden et al., 2019; Gros et al., 2019), we systematically study the impact of the above data preprocessing and training strategies to develop a robust deep learning-based method for the automatic segmentation of MS lesions in axial T2w scans covering the entire spine. Specifically, we:

1. Use a region-based approach to simultaneously segment MS lesions and the spinal cord. We improve upon the performance of the existing tools (Bédard et al., 2023; Gros et al., 2019), particularly in the lower-thoracic and lumbar spines.
2. Evaluate preprocessing methods such as stitching individual chunks versus training on raw chunks and the effects of straightening versus no straightening.
3. Compare (patch-wise) 3D versus (slice-by-slice) 2D kernel training strategies for spinal cord lesion segmentation with convolutional neural networks.
4. Investigate generalizability across various sites and imaging protocols for spinal cord lesion segmentation.

## 2 Materials and Methods

### 2.1 MRI Data and Ground Truth

MRI data were retrospectively collected from four different sites, encompassing both cross-sectional and longitudinal datasets, with partial and full spinal cord coverage, from a total of 582 patients, following internal review board approval and performed in accordance with the Declaration of Helsinki.

The longitudinal dataset consisted of 317 patients acquired at a single site, namely the Klinikum Rechts der Isar, Technical University of Munich, Munich, Germany (referred to as the *TUM* dataset). Each patient underwent two MRI examinations (sessions), with each session containing 3 individual chunks of highly anisotropic axial T2-weighted scans covering the cervical, thoracic, and lumbar spines, respectively. The chunks are numbered 1-3 in the cranio-caudal direction for each spinal region. Participants for this site spanned a large variety of clinical phenotypes and conditions, namely, clinically isolated syndrome (CIS; *n* = 11), radiologically isolated syndrome (RIS; *n* = 1), primary progressive MS (PPMS; *n* = 11), secondary progressive MS (SPMS; *n* = 14), relapsing-remitting MS (RRMS; *n* = 277) and unknown (n=3). The dataset comprises patients with spinal cord lesions and those without lesions at one or both longitudinal time points. The mean time interval between baseline and follow-up scan (in years) was 3.1 ± 2.6, with an average age of 36.1 ± 10.4 at baseline. Of the 317 patients, the female-/male- ratio was 209/108.

The ground-truth (GT) spinal cord and lesion masks were manually annotated by two experienced raters (SR and RW). Annotations were guided by an iterative pre-labeling scheme, which involved an initial training phase where a segmentation network was trained on a manually labeled subset of the cohort. Intermediate segmentations were obtained from this trained network followed by manual corrections by raters SR and RW. This pre-labeling approach significantly expedited the annotation process, enabling the inclusion of a large number of patients and whole spine scans, which would have been practically infeasible with manual annotation alone.

Starting from the raw chunks of the individual spinal regions in the native space (*Chunks Native*), Figure 1A shows the dataset characteristics of the different preprocessed variants of the TUM dataset obtained after stitching and straightening the chunks. More details are given in Section 2.3 and Figure 2.

**Figure 1:**
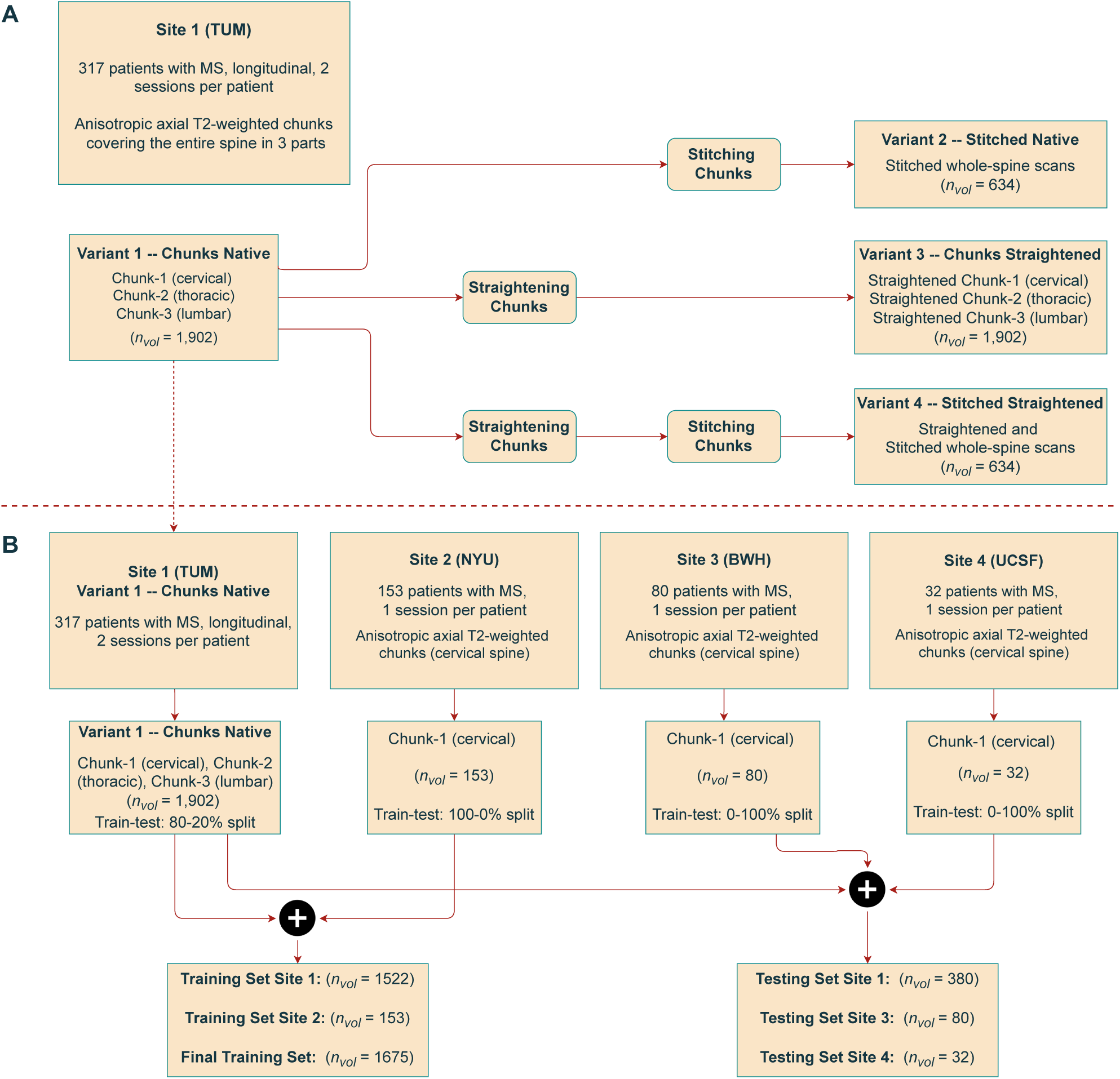
Study flowchart. (A): Characteristics of the TUM dataset along with its preprocessed variants. (B): Characteristics of data from different sites and the respective train/test splits. *n* denotes the number of patients and *n_vol_* denotes the total number of images.

**Figure 2:**
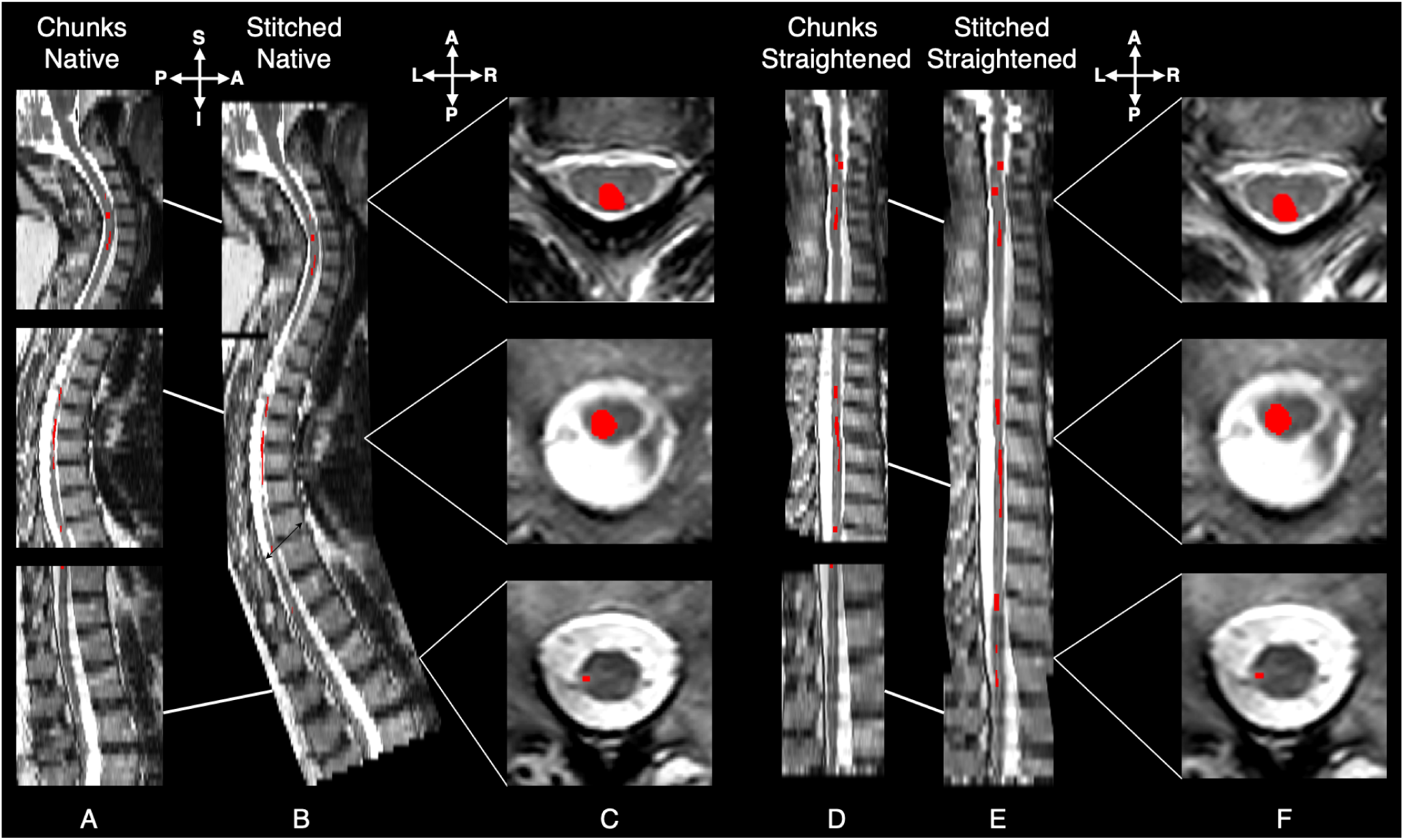
Illustration of the dataset versions and respective lesion appearance. Each image shows the middle sagittal slice of the axial T2w sequence from a representative subject in the dataset. Lesions, highlighted in red, represent the maximum intensity projection of the lesion mask on the sagittal slice, allowing visualization of all lesions along the whole spinal cord. **(A)**: Middle sagittal slice of a stack of three axial T2w acquisitions covering the cervical, thoracic, and lumbar regions of the spine. **(B)**: Stitched whole-spine scan derived from the respective chunks. **(C)**: Axial slice of lesions in each chunk. **(D)**: Straightened version of the individual chunks, where the curvature of the cord was removed using `sct_straighten_spinalcord`. **(E)**: Stitched configuration of the previously straightened chunks. **(F)**: Axial slice of lesions in the straightened chunk. In both stitched and straightened variants, the lesion characteristics remain consistent, indicating a (desired) minimal impact on lesion morphology. evaluate how the different formats of the input scans affected the downstream lesion segmentation performance.

For the cross-sectional datasets, 153 patients were scanned at the NYU Langone Medical Center, New York, USA, 80 subjects were scanned at the Brigham and Women’s Hospital, Harvard Medical School, Boston, USA, and 32 were scanned at the Zuckerberg San Francisco General Hospital, San Francisco, California, USA. The scans from all three sites primarily covered the cervical or cervico-thoracic spines with only either of the chunks for each patient. The lesion masks were annotated manually by raters at the respective sites. The spinal cord masks were generated with sct deepseg sc using Spinal Cord Toolbox (De Leener, Lévy, et al., 2017) followed by manual corrections wherever necessary. Clinical data were not available for the MS patients across the sites. Following our experiments with the chunks from the TUM dataset, the scans from these three sites were primarily used to improve and test the segmentation model’s generalization capabilities. Figure 1B shows how the data from different sites are combined for developing the lesion segmentation model. Table 1 shows a detailed overview of the image characteristics and acquisition parameters from all sites in this study.

**Table 1:** Overview of image characteristics and acquisition parameters from each site. TR and TE represent the median repetition and echo times, respectively.

| Site | #Scans | Manufacturer | TR (ms)<br>(median) | TE (ms)<br>(median) | Resolution (mm<sup>3</sup>) |
| --- | --- | --- | --- | --- | --- |
| Klinikum Rechts der Isar,<br>Technical University of Munich,<br>Munich, Germany | 1999 | Siemens 1.5T ($n = 14$) | 6065 | 104 | min: $0.19 \times 0.19 \times 1.95$<br>max: $0.78 \times 0.78 \times 6.83$ |
| | | Siemens 3T ($n = 290$) | 7070 | 107 | |
| | | Philips 1.5T ($n = 5$) | 3034 | 100 | |
| | | Philips 3T ($n = 1687$) | 4435 | 90 | |
| | | GE 1.5T ($n = 3$) | 6280 | 114 | |
| NYU Langone Medical Center,<br>New York, USA | 209 | Siemens 3T ($n = 209$) | 4000 | 107 | min: $0.56 \times 0.56 \times 3.3$<br>max: $0.78 \times 0.78 \times 11.91$ |
| Brigham and Women’s Hospital,<br>Harvard Medical School,<br>Boston, USA | 80 | Siemens 3T ($n = 80$) | 5070 | 101 | min: $0.56 \times 0.56 \times 3$<br>max: $0.7 \times 0.7 \times 3$ |
| Zuckerberg San Francisco<br>General Hospital, San Francisco,<br>California, USA | 32 | GE 3T ($n = 32$) | 3516 | 72 | min: $0.29 \times 0.29 \times 3$<br>max: $0.7 \times 0.7 \times 6$ |

### 2.2 Preprocessing framework

#### 2.2.1 Stitching

In clinical practice, full spinal cord scans are impractical due to long acquisition times. An alternative is to *stitch* individual segments into whole-spine scans. In this paper, we refer to *stitching* as the process of aligning and merging multiple image segments or chunks, in this case, axial T2-weighted (T2w) scans acquired during the same session, into a single image. It must be noted that chunks may also be registered to the template individually, however, it is preferred to directly register the stitched image as this simplifies the computation of lesion morphometrics by eliminating the need to reconcile overlapping regions or risk double-counting that can occur when multiple chunks are processed separately (Graf, Möller, et al., 2024).

To compare whether the model benefits from the extended context around the spinal cord in stitched whole-scans, we employed the stitching algorithm implemented in (Graf, Platzek, et al., 2024; Lavdas et al., 2019). This algorithm has four main steps: (i) the corner points are computed for each chunk, (ii) using the affine matrix of each chunk, corner points are rotated so as find a minimum bounding box that fits all the chunks, resulting in optimal rotation and spacing parameters, (iii) the chunks are resampled to the new (common) image space and occupancy maps containing overlapping regions between chunks are stored, (iv) the resampled chunks are blended using ramp-based interpolation with a weighted averaging function to ensure smooth transitions between chunks. Manual corrections are applied to the stitched mask, if required, to reduce interpolation artifacts. Figure 2 (A-C) shows the result of the stitching algorithm the subsequent appearance of lesions in the whole-spine scan.

#### 2.2.2 Straightening

In the analysis of brain MS lesions, MRI scans of the brain are typically registered to the MNI anatomical template using rigid registration (Carass et al., 2017a; Wiltgen et al., 2024). Following this approach of analyzing the lesions in a template space, one would have to register the individual chunks to, for e.g., the PAM50 template (De Leener et al., 2018). As it is difficult to obtain the vertebral levels in axial scans with thick sagittal slices, we opted for a simpler approach involving *straightening* of the spinal cord (De Leener, Mangeat, et al., 2017). Straightening essentially eliminates all curvature-related variance in the spinal cord, offering a simpler alternative to template-based registration and facilitating cohort-level analysis. We hypothesize that eliminating patient-level differences arising from the spinal cord curvature may improve automatic lesion segmentation. The straightening procedure using Spinal Cord Toolbox (SCT) (De Leener, Lévy, et al., 2017) is described as follows: Given an axial T2w scan (chunk or stitched), we applied sct straighten spinalcord using the spinal cord segmentation mask as reference to obtain the straightened cord along with the warping field (warp curve2straight). Then, the output warping field was applied using sct apply transfo with linear interpolation to straighten both the GT spinal cord and the lesion masks. Lastly, both the straightened GT masks were binarized using a threshold of 0.5. Figure 2 (D-F) shows the straightened individual chunks, straightened whole-spine scans and the resulting appearance of lesions across spinal regions, with a reduced field of view resulting from the straightening algorithm.

### 2.3 Experimental Design

Experiments were designed following two major themes: (i) investigating the effects of data preprocessing (namely, stitching and straightening the chunks) on lesion segmentation using the TUM dataset, and building on this (ii) leveraging data from additional sites to obtain a robust and generalizable segmentation model for MS lesions in axial T2w scans.

Regarding the first theme, we created four versions of the *TUM* training dataset (see Figure 1A) to evaluate how the different formats of the input scans affected the downstream lesion segmentation performance.

1. **chunks native**, the original dataset containing the individual chunks of axial T2w scans corresponding to cervical, thoracic, and lumbar regions of the spine,
2. **stitched native**, consisting of a single scan per patient with the individual cervical, thoracic and lumbar chunks *stitched* together to form the whole spinal cord,
3. **chunks straightened**, where the individual chunks in their native space are straightened using the spinal cord segmentation masks and the model is trained in the straightened image space, and,
4. **stitched straightened**, where the stitched images in their native space are straightened using the whole-spine cord segmentation masks.

To explore the effect of spatial context in terms of training on individual slices or 3D patches, models with both 2D and 3D kernels were trained on each of the four dataset variants, resulting in 8 models in total. Only anisotropic, axially acquired T2-weighted scans were used in our experiments. Furthermore, because of the evolution in the appearance of lesions between sessions, the images were treated as independent inputs for training the segmentation model. The TUM dataset was split *patient-wise* according to 80-20% train/test ratio to ensure that the chunks for a given patient belonged either to the training set or testing set but not to both. For the **chunks native** dataset, this resulted in a total of 1522 individual chunks from 254 patients for training/validation and 380 chunks from 63 patients. For the **stitched native** dataset, this translated into 508 stitched whole-spine scans from 254 patients in the training/validation set and 126 scans from 63 patients in the testing set.

As for the second theme focusing on robustness and generalization, all 153 patients’ chunks from the NYU site were added to the training set while keeping the BWH and UCSF sites unseen, in order to test the model’s generalization to out-of-distribution data. Such a training and evaluation approach partly overlaps with a lifelong learning scenario where the segmentation model is *continually* enriched with data from new sites by adding them to current training set and testing on data from unseen sites (Naga Karthik et al., 2022). Figure 3 shows the distribution of lesions in terms of the lesion volume across train and test splits for each site.

**Figure 3:**
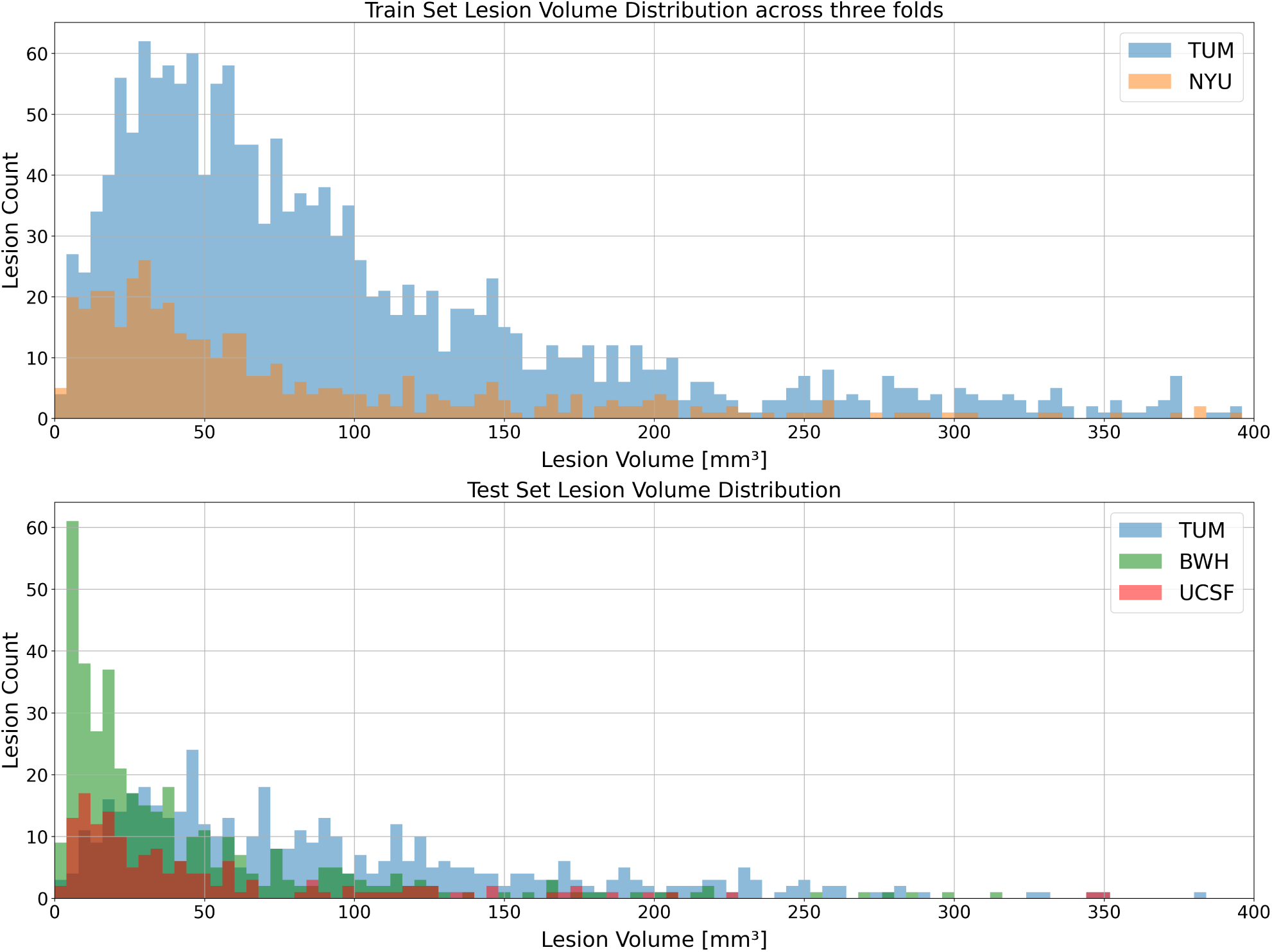
Lesion Distribution Plot across sites in train and test sets. Outliers with lesion volumes exceeding 400 *mm*^3^ were excluded for visualization purposes only (not for model training). All sites and sets follow a similar distribution of lesion count over size, validating a meaningful train/test split for the experiments and training of the final model.

## 2.4 Training Protocol

The continuing dominance of nnUNet (Isensee et al., 2021, 2024) across several open-source segmentation challenges has shown that a well-tuned convolutional neural network (CNN) architecture is robust and continues to achieve state-of-the-art results over novel transformer-based architectures (Hatamizadeh et al., 2021; Karthik et al., 2024; Ma et al., 2024). Based on this rationale, we used nnUNetV2 (Isensee et al., 2021) as the segmentation framework. Specifically, we used nnUNet’s region-based training strategy, where the model simultaneously segments the spinal cord and lesions. The rationale for going with this approach is that combining the label of a larger object (i.e. the spinal cord) with a smaller one (i.e. the lesion) might help obtain better gradients for effective learning early on during the training. Moreover, framing spinal cord and lesion segmentation as a hierarchical segmentation problem provides the model with the perspective that lesions may only occur within the spinal cord, potentially offering a better guidance mechanism to avoid segmenting imaging artifacts outside the spinal cord, given the small sizes of lesions (Isensee et al., 2019). While region-based training requires both spinal cord and lesion masks as inputs, the rise of robust spinal cord segmentation tools (Bédard et al., 2023; Gros et al., 2019; Naga Karthik et al., 2024) across various MRI contrasts and pathologies enables obtaining spinal cord segmentation masks without major manual intervention.

Default data augmentation transforms applied by nnUNet were used, namely, random rotation, scaling, mirroring, Gaussian noise addition, Gaussian blurring, adjusting image brightness and contrast, low-resolution simulation, and gamma transformation. All scans were converted to RPI orientation and preprocessed using Z-score normalization. For each of the four preprocessed datasets, 2D and 3D models were trained using three-fold cross validation resulting in a total of 24 models. All models were trained for 1000 epochs, with a batch size of 2, using the stochastic gradient descent optimizer with a polynomial learning rate scheduler.

While nnUNet is known for automatically configuring the patch sizes of the images used during training based on the hardware capacity, its default patch sizes could be sub-optimal depending on the size of the input images. Specifically, in the case of the *stitched* datasets, the automatically-configured patch sizes for 3D models were defined to be half the size of the median shape of the images from the training set. This means that the model effectively received only half of the total length of the (stitched) spinal cord in the S-I plane as a 3D input patch during training. To evaluate whether fitting the whole context of the cord within a single input patch would improve the segmentation performance, we also trained a model on a patch size covering the entire S-I plane, meaning that the model has access to the whole context of the spinal cord in the sagittal plane.

All segmentation networks were trained with nnUNet v2.4.1 using Python 3.10.13 and Pytorch 2.2.1. Training both 2D and 3D models on any given dataset variant took approximately 1 day on a single NVIDIA A100 GPU with 40 GB memory.

## 2.5 Evaluation Protocol and Metrics

### Spinal cord segmentation

We compared our models on the TUM test set with three other open-source methods available in SCT: (i) `sct_propseg` (De Leener et al., 2014), (ii) sc deepseg sc (Gros et al., 2019), and (iii) the recently proposed contrast-agnostic segmentation model (Bédard et al., 2023). The test split was done at the patient-level (and not at the chunks-level), ensuring that the axial scans of a particular patient strictly belonged only to the testing set, resulting in a total of 126 patients. The performance of these models was then evaluated independently on each of the three chunks (i.e. cervical, thoracic and lumbar) from the TUM dataset.

### Lesion segmentation

We created three test sets: the TUM (*n* = 63) and the chunks from sites, BWH (*n* = 80), and UCSF (*n* = 32) (which were kept unseen during training) to evaluate the model’s performance on out-of-distribution data. First, to determine the dataset variant and training strategy that leads to best performance, 8 models (4 dataset variants, 2 models each after combining the results from 3 folds) were compared. Then, the best model out of the 8, the model trained on chunks from two sites, along with `sct_deepseg_lesion` (Gros et al., 2019), were evaluated on the TUM, BWH, and UCSF test sets.

### Metrics

While the `ANIMA` toolbox (Commowick et al., 2021), specifically `animaSegPerfAnalyzer`, is widely used for the evaluation of brain MS lesion segmentation models (Commowick et al., 2021), it fails to output appropriate metrics in special cases where the ground-truth mask contains no lesions, *irrespective* of whether the model predicts false positive lesions or not. Skipping such cases in the evaluation of spinal cord MS lesions would result in (biased) higher scores for the metrics by not accounting for the false positive rate of the segmentation models. To address these limitations, we used the open-source `MetricsReloaded package` (Maier-Hein et al., 2024; Reinke et al., 2024), designed to mitigate several shortcomings of existing open-source toolboxes. In the context of this study, when the GT and the automatic prediction are both empty (i.e. contain no lesions), the voxel-wise Dice is set to 1, indicating the model’s ability to learn correctly. When either of them is non-empty, the respective false-positive and false-negative scores are computed by formal definition. As for metrics, for spinal cord segmentation, we computed the voxel-wise Dice score, normalized surface distance (NSD), and relative volume error (RVE). For lesion segmentation, we reported the voxel-wise Dice score, NSD, and average absolute difference of the lesion count between the number of predicted lesions and GT lesions, and lesion-wise F_1_ score, where a prediction was considered true positive if it showed at least a 10% overlap with the GT lesion.

To ensure a fair comparison between the models trained on the four variants of the TUM dataset, we used the following post-processing strategies before computing the metrics:

1. **Chunks Native**: For each patient, the 3 chunks corresponding to cervical, thoracic and lumbar spines were stacked together to obtain a single prediction for a given patient (instead of having 3 lesion prediction masks per patient). As the sizes of the chunks differed depending on the spinal region, an appropriate amount of padding was applied to the chunks to enable stacking. Figure 4 illustrates our concept of stacking and algorithm 1 describes it in practice.
2. **Stitched Native**: No post-processing was done in this case, as we already have a single prediction per patient.
3. **Chunks Straightened**: To ensure that all predictions are evaluated in the same space, the straightened chunks were brought back to the image space by applying the inverse warping field. This was followed by stacking the chunks (now in the native space) to obtain a single prediction per patient.
4. **Stitched Straightened**: Similar to the straightening of chunks, the straightened stitched images were transformed back to the native space by applying the inverse warping field.

**Figure 4:**
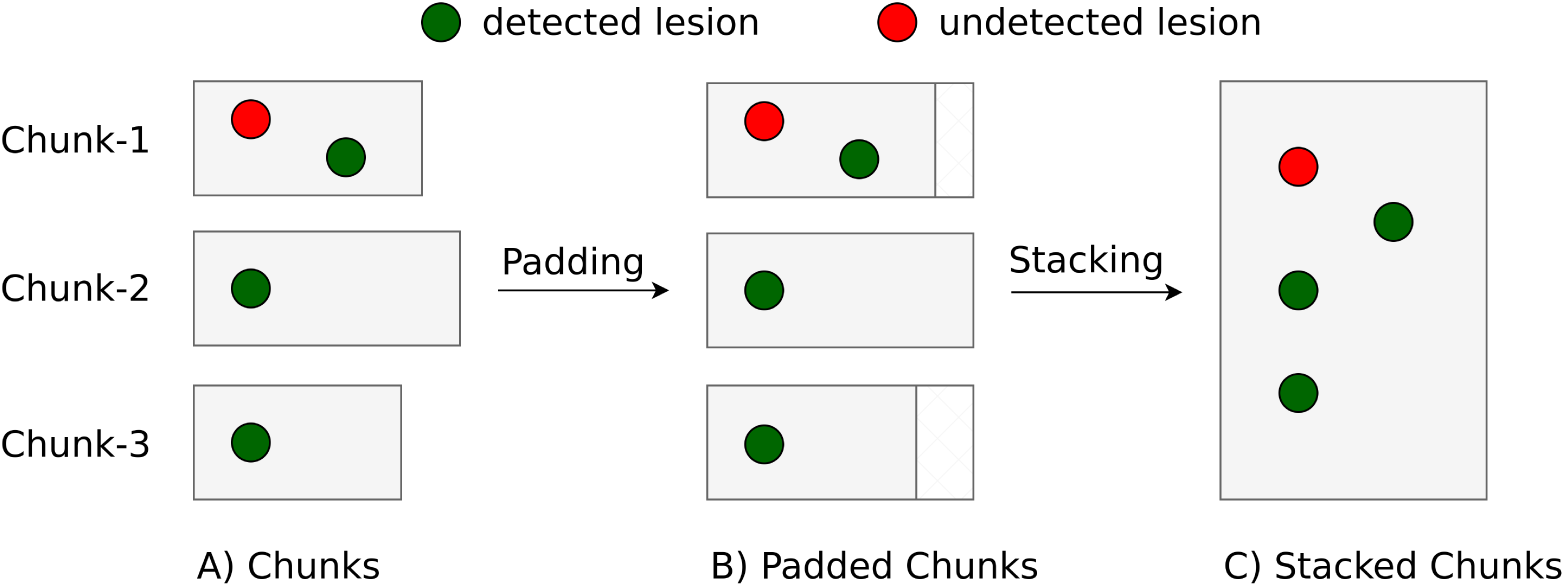
Toy example illustrating the difference in computing the Recall score with and without stacking. When computed *chunk-wise* (A), 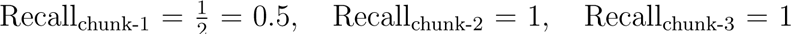 the average F1 score across chunks is calculated as: 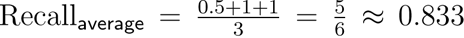. In contrast, the Recall score on stacked chunks (C) is given by: 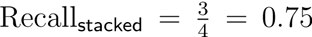. This illustrates that averaging the Recall scores computed individual chunks yields a higher value than computing the same on stacked chunks. Although the correct computation of metrics can be achieved without stacking through a straightforward mathematical approach, we opt for stacking as it simplifies the handling of prediction and GT masks and the metrics computations with the `MetricsReloaded toolbox`.

Given the difference in the magnitude of F_1_-scores computed on individual and stacked chunks, all metrics described in Section 3 were computed on stacked chunks, the models were trained on chunks.

The process of stitching chunks can lead to the interpolation of lesions in overlapping areas, while the process of stacking may result in double-counting these lesions. To assess whether this has a significant impact on our dataset, we analyzed our TUM test set. Among a total of 126 stitched scans (or respectively 378 chunked scans), we observed 450 lesions in the chunked dataset and 448 in the stitched dataset, a difference that can be attributed to a single patient. Given the minimal frequency and negligible impact of this discrepancy, we decided not to account for this effect in our analysis.

**Algorithm 1:**
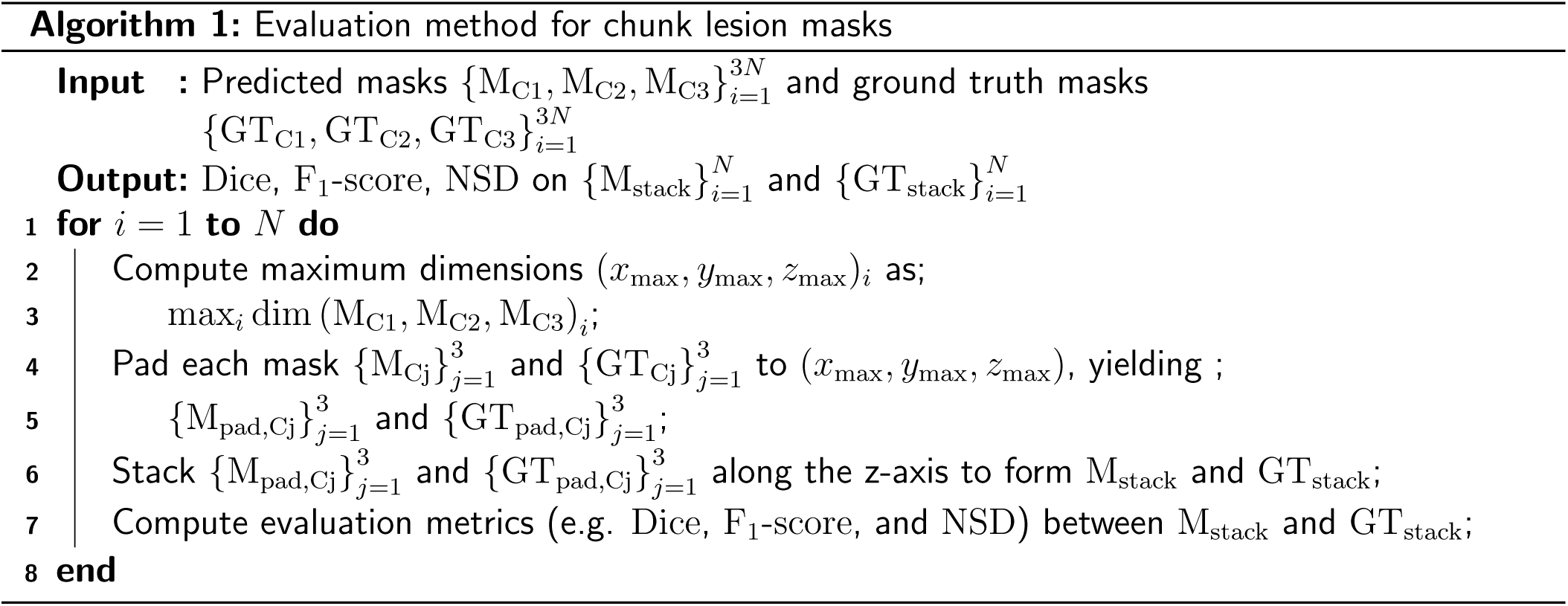
Evaluation method for chunk lesion masks

### Statistical Analysis

Statistical analysis was performed using the SciPy Python library v1.9.1 (Virtanen et al., 2020). Data normality was tested using D’Agostino and Pearson’s normality test. Group-wise comparison between the 2D and 3D models for each of the four variants of the TUM dataset was performed using the non-parametric Kruskal-Wallis H-test. Post hoc pairwise test for multiple comparisons was performed using Dunn’s test with holm correction.

## 3 Results

This section compares the results of our proposed model for spinal cord segmentation against various existing methods (Section 3.1). Next, we examine how spinal cord straightening affects the performance of the lesion segmentation model (Section 3.2). We then demonstrate the model’s capability to detect lesions of varying sizes and evaluate its performance in detecting lesions across different chunks (Section 3.3). Finally, we identify the best model based on these findings and enhance it further by incorporating additional data from external sites (Section 3.4).

### 3.1 Spinal cord segmentation

Figure 5 compares the performance of 2D/3D models trained on chunks/stitched images respectively with state-of-the-art methods for spinal cord segmentation evaluated independently across different spinal regions (cervical, thoracic and lumbar). Interestingly, spinal cord segmentations from `sct_propseg` and `sct_deepseg_sc` vary drastically across chunks, with the lowest performance on chunk-3 covering thoracolumbar levels. The `contrast-agnostic` v2.5^2^ model (Bédard et al., 2023) reduces the gap in segmentation accuracy across chunks with a notable improvement in the lumbar spine segmentation. However, the models trained with TUM data (chunk-wise or stitched) perform significantly better than other methods for our test set.

**Figure 5:**
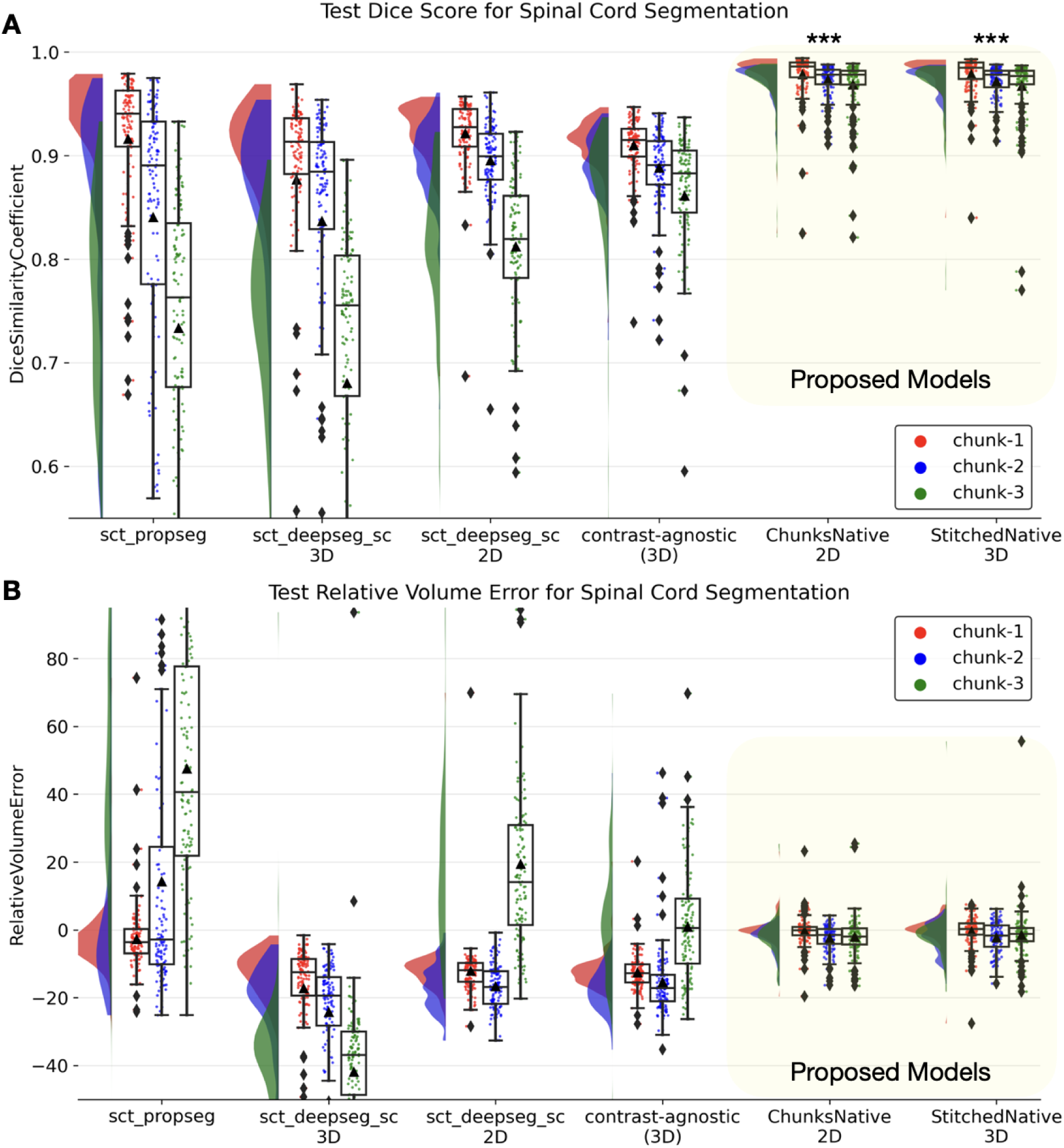
Raincloud plots comparing the (**A**) Dice scores (best: 1; worst: 0) and (**B**) relative volume error (in %, best: 0%) across various spinal cord segmentation methods evaluated independently on the three chunks. The proposed Chunks and Stitched models significantly outperform the rest of the methods across all chunks. *** *P* < .001 (group-wise Kruskal-Wallis H-test followed by post-hoc pairwise Dunn’s test for multiple comparisons). Statistically significant pair-wise differences were found throughout when comparing each existing method with each of the proposed models.

### 3.2 Impact of cord curvature and spatial context

In this section, we compare the lesion segmentation performance across the different variants of the TUM dataset described in Section 2.3. Figure 6 shows multiple raincloud plots comparing the lesion segmentation performance of models trained on individual axial slices (2D) and three-dimensional image patches (3D). Training a 2D model resulted in a higher Dice and F_1_-score compared to the 3D models in all datasets except for the *stitched native* variant.

**Figure 6:**
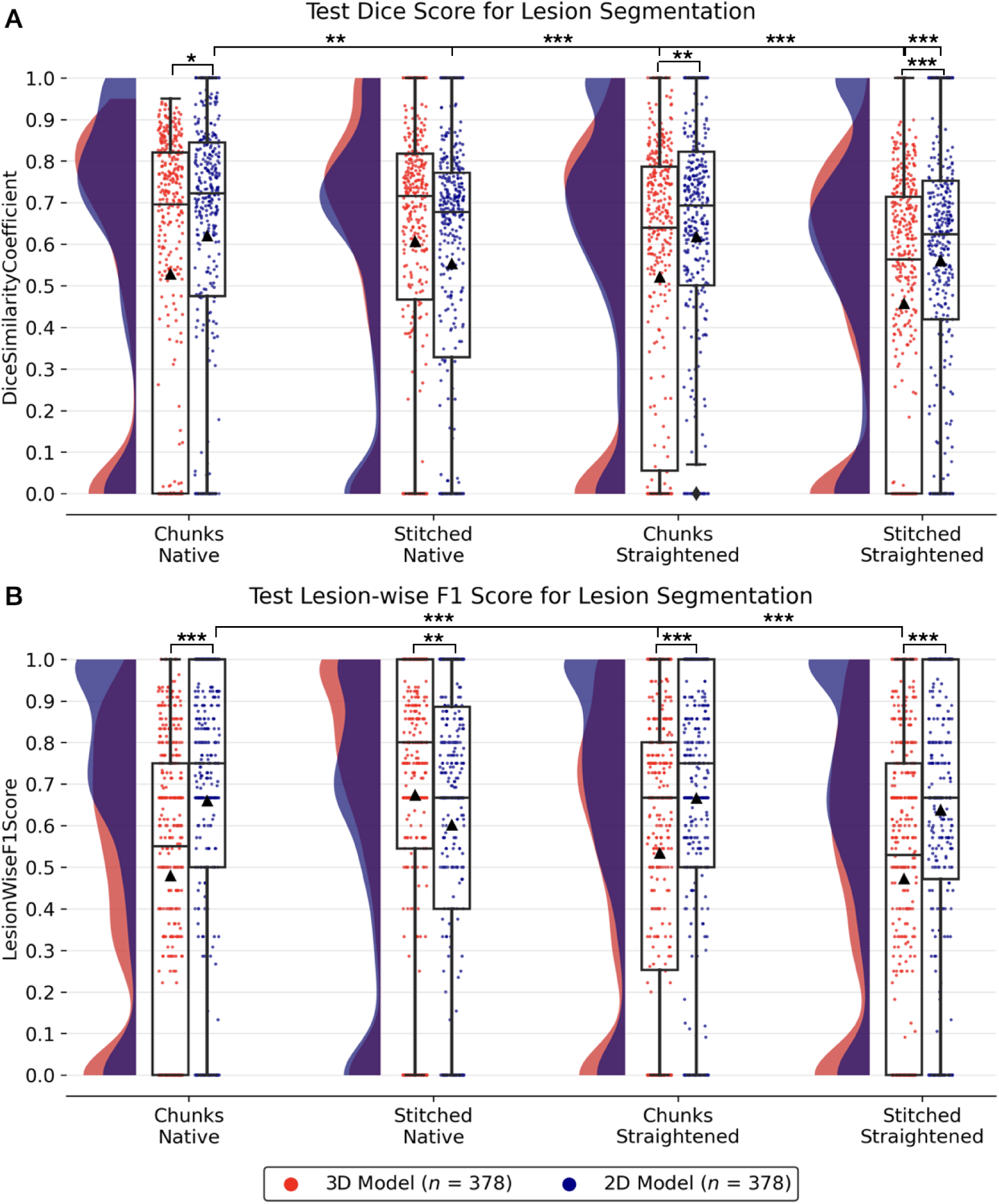
Raincloud plots comparing test lesion segmentation performance in terms of voxel-wise Dice (A) and lesion-wise F1-score (B) across 2D/3D models trained on 4 variants of the TUM dataset. Scatter points represent the model prediction on a single image from a given fold. Distributions illustrate the spread of predictions and ▴ represents the mean value over 3 folds. * *P < .*05, ** *P < .*01, *** *P < .*001 (Group-wise Kruskal-Wallis H-test followed by post-hoc Dunn’s test adjusted for multiple comparisons with Holm correction).

As mentioned in Section 2.4, along with default patch sizes defined by nnUNet, we also experimented with modified patch sizes covering the entire length of the cord within a single input patch to see if extended patch sizes would improve segmentation performance. Interestingly, we found no statistically significant differences between the 3D models trained with default patch sizes and the models trained with a full S-I coverage. Hence, we proceeded with the default patch sizes for all models.

Considering the role of spinal cord curvature in lesion segmentation, we observed that the models trained on straightened cords performed slightly worse than the models trained in the native space of the input scans. Specifically, in 2D, a median Dice score of 0.72 (*chunks native*) vs. 0.69 (*chunks straightened*) and, in 3D, a median Dice of 0.72 (*stitched native*) vs. 0.53 (*stitched straightened*). This is also apparent from the wider distribution of the Dice scores of predictions (i.e. scatter points) in Figure 6A. In a between-dataset pairwise comparison of Dice scores, the 2D *chunks native* model significantly outperformed 2D *stitched native* (**^**^** *P < .*01), 3D *chunks straightened* (**^***^** *P < .*001), and 2D/3D *stitched straightened* models (**^***^** *P < .*001). Likewise, for lesion-wise F_1_-scores, the 2D *chunks native* model showed significant differences between 3D *chunks straightened* (**^***^***P* < .001), and 3D *stitched straightened* models (**^***^** *P* < .001).

### 3.3 Lesion distribution, frequency and detection across chunks

Considering the median Dice and F_1_-scores, the *Chunks Native* 2D model ranks higher than the straightened counterparts and is on par with the models trained on stitched scans. In other words, training directly on chunks is a simple strategy that does not perform worse than the stitched models *without* the need for any stitching procedure. As this presents a simple and scalable solution, we proceeded with the 2D model and present the results from our subsequent experiments evaluating lesion detection in this section.

Figure 7 shows the lesion detection rates across various categories of lesions sizes. The performance of the model increases with increasing lesion sizes even detecting small lesions between 10-50 mm^3^ at a 60% rate. We observed that lesion detection becomes challenging as lesions get even smaller.

**Figure 7:**
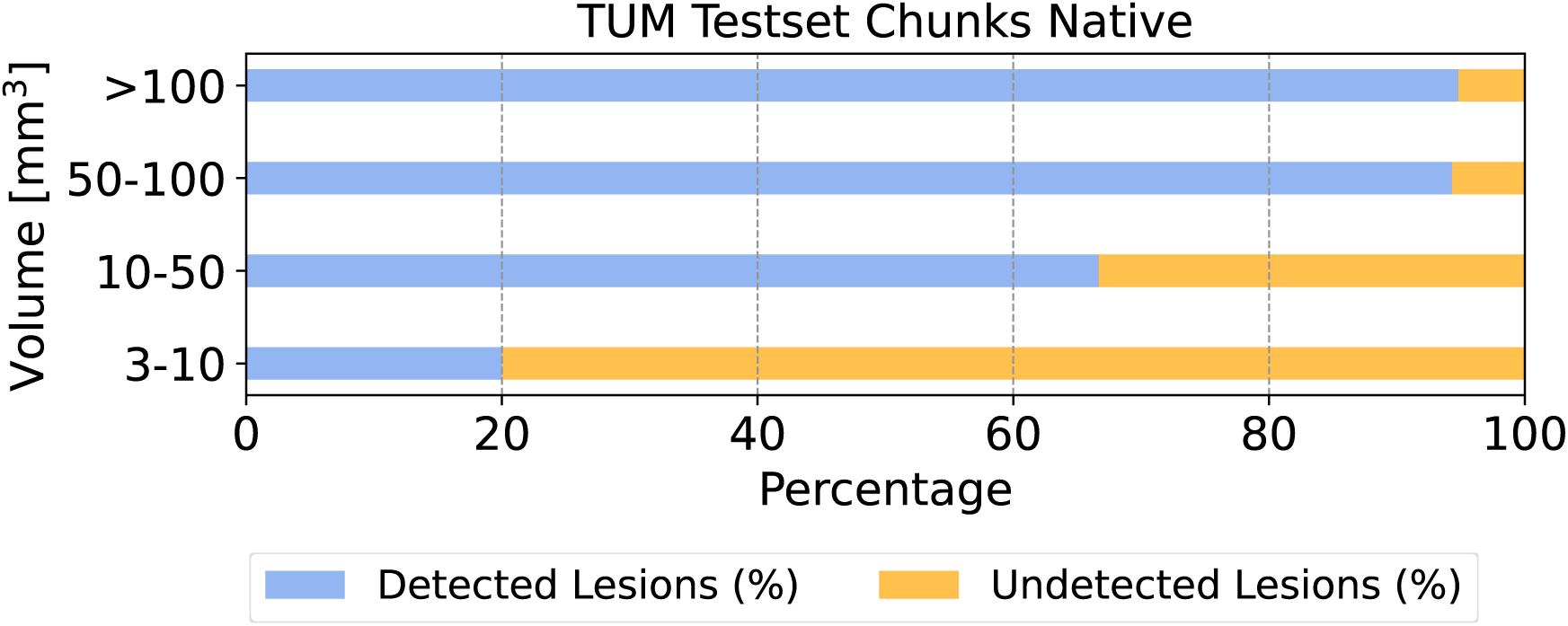
Lesion detection rates across four volumetric lesion categories for the native chunks dataset using the 2D model. While the model struggles to segment very small lesions, which are often difficult to distinguish from image artifacts and contribute to high inter-rater variability, it demonstrates high detection rates for medium and large lesions.

To evaluate whether our model is consistent in detecting lesions occurring at various spinal cord regions, we plotted the distribution of lesion sizes and the corresponding lesion detection rate across spinal cord chunks in Figure 8. As expected, we observed a decreasing trend in the frequency of lesions going from the cervical to thoracolumbar spine. Our model achieved similar lesion detection rates in cervical and cervicothoracic spines (left and middle), with a slight decrease in the thoracolumbar spine (right).

**Figure 8:**
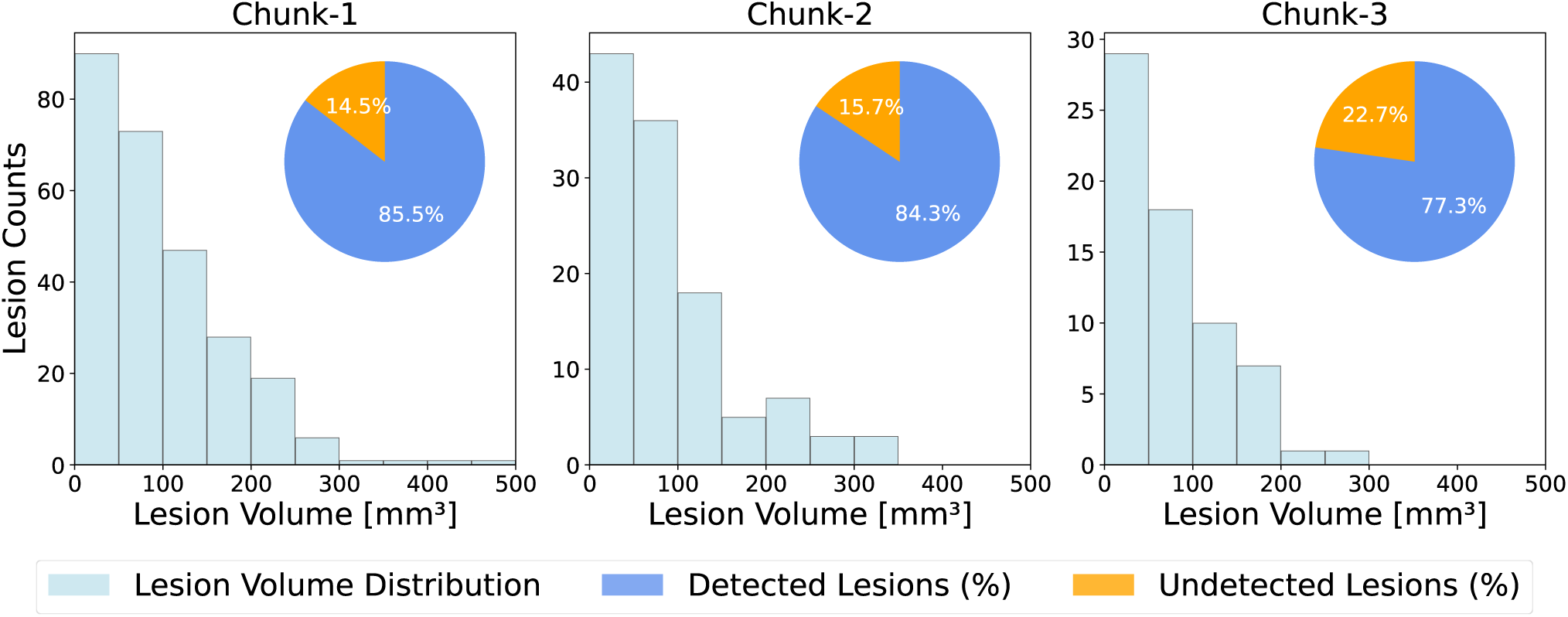
Histograms of spinal cord lesions across all TUM test set chunks, showing higher lesion prevalence in cervical spine (chunk-1), but similar trends in terms of lesion volume distributions across chunks (2-3). Note the different scaling of the y-axes. Lesion detection rates across the different chunks for the TUM test set slightly decrease when traversing the spine cranio-caudally.

### 3.4 Generalizability across sites and protocols

We have now established that the model trained on chunks performs at least on par as the stitched models *without any explicit preprocessing* (namely, stitching and straightening) with similar lesion detection rates across chunks. Following the experimental design, we added axial T2w chunks from an external site (NYU) and evaluate whether the performance of the model improves after training on two sites (TUM and NYU). We tested the model in two ways: (i) on an *in-distribution* test set (i.e. TUM) to evaluate whether the model’s performance has drifted after the addition of an external site, and (ii) on unseen (out-of-distribution) data from two additional sites (BWH and UCSF), acquired using different scanners and protocols unseen during training.

Table 2 and Figure 9 compare three models qualitatively and quantitatively: (i) sct deepseg lesion, the current state-of-the-art, (ii) the model trained on TUM dataset’s chunks (i.e. single site), and (iii) the model trained on two sites. As data from BWH and UCSF are acquired at different sites and with acquisition parameters, the results on these sites show the model’s robustness to out-of-distribution data. Both variants of the models trained on chunks performed significantly better than sct deepseg lesion (Gros et al., 2019), which tends to miss lesions (shown with yellow arrows) resulting in higher false negatives. Importantly, adding data from the NYU site did not degrade the model’s performance on the original TUM test set, rather, it showed a slight improvement across all metrics. As for the evaluation metrics on unseen sites, the model trained on two sites performed better than the single site model, although no statistically significant difference was found.

**Figure 9:**
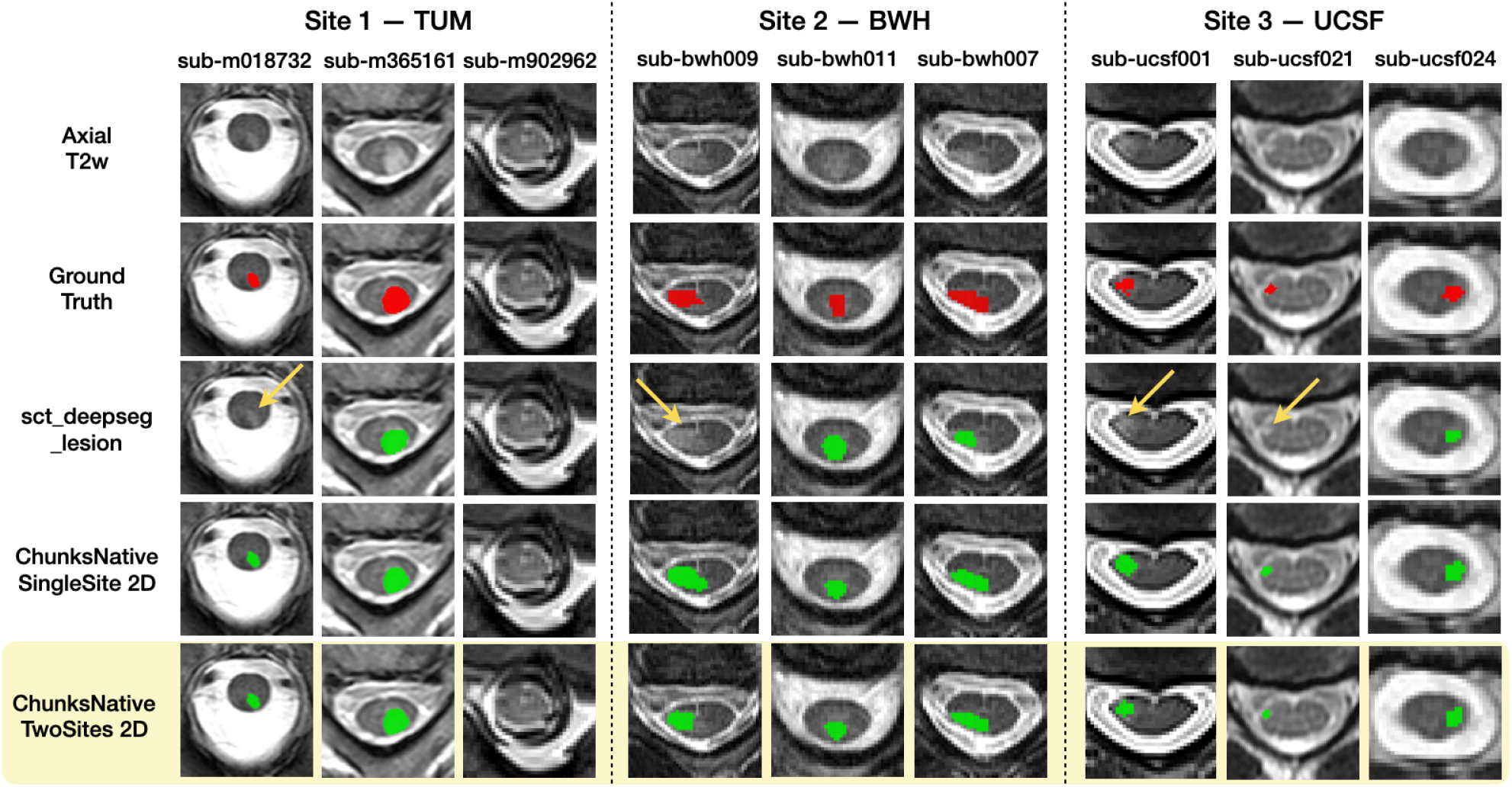
Qualitative comparison of sct deepseg lesion, *ChunksNative SingleSite* and *ChunksNative TwoSites* 2D models across three patients from each test site. `sct_deepseg_lesion demonstrates` low sensitivity to MS lesions, missing to segment lesions in a few patients (yellow arrows).

**Table 2:** Quantitative comparison of the models trained on chunks on in-distribution (TUM) and out-of-distribution test sites (BWH and UCSF). Number of images for each test site is given in brackets.

| Test Site | Metric | sct_deepseg<br>_lesion | ChunksNative<br>SingleSite | ChunksNative<br>TwoSites |
| --- | --- | --- | --- | --- |
| **TUM**<br>($n = 126$) | Dice<sup>↑</sup> | $0.33 \pm 0.31$ | $0.60 \pm 0.34$ | **$0.62 \pm 0.33$**<sup>†</sup> |
| | NSD<sup>↓</sup> | $0.37 \pm 0.31$ | $0.65 \pm 0.37$ | **$0.67 \pm 0.36$** |
| | F1ScoreL<sup>↑</sup> | $0.42 \pm 0.34$ | $0.64 \pm 0.36$ | **$0.66 \pm 0.34$**<sup>†</sup> |
| | $\vert n_{\text{refL}} - n_{\text{predL}}\vert $<sup>↓</sup> | $1.93 \pm 1.87$ | $1.86 \pm 1.89$ | **$1.62 \pm 1.81$** |
| **BWH**<br>($n = 80$) | Dice<sup>↑</sup> | $0.28 \pm 0.30$ | $0.45 \pm 0.26$ | **$0.51 \pm 0.26$**<sup>†</sup> |
| | NSD<sup>↓</sup> | $0.34 \pm 0.31$ | $0.60 \pm 0.32$ | **$0.65 \pm 0.30$** |
| | F1ScoreL<sup>↑</sup> | $0.40 \pm 0.37$ | $0.55 \pm 0.32$ | **$0.62 \pm 0.32$**<sup>†</sup> |
| | $\vert n_{\text{refL}} - n_{\text{predL}}\vert $<sup>↓</sup> | $3.45 \pm 4.22$ | $3.27 \pm 3.85$ | **$3.00 \pm 3.82$** |
| **UCSF**<br>($n = 32$) | Dice<sup>↑</sup> | $0.15 \pm 0.16$ | $0.52 \pm 0.18$ | **$0.53 \pm 0.18$**<sup>†</sup> |
| | NSD<sup>↓</sup> | $0.24 \pm 0.25$ | $0.69 \pm 0.22$ | **$0.71 \pm 0.21$** |
| | F1ScoreL<sup>↑</sup> | $0.25 \pm 0.26$ | $0.68 \pm 0.23$ | **$0.69 \pm 0.23$**<sup>†</sup> |
| | $\vert n_{\text{refL}} - n_{\text{predL}}\vert $<sup>↓</sup> | $3.22 \pm 2.88$ | $2.22 \pm 1.81$ | **$1.69 \pm 2.01$** |

Note: Data are means ± standard deviations. The best values for NSD and $|n_{\text{refL}} - n_{\text{predL}}|$ are 0.0 and 1.0 for Dice and F1ScoreL. Bold represents best performance.
<sup>†</sup> Statistically significant compared to DeepSegLesion ( $P < .001$ )

## 4 Discussion

In this study, we developed an automatic tool for the joint segmentation of the spinal cord and MS lesions in axial T2w MRI scans using a region-based training approach. Starting with a comprehensive comparison between chunks, stitched, and straightened variants of our dataset, we showed that training a 2D model directly on the raw chunks in the native space achieved the best performance, presenting a simple and scalable solution. With respect to spinal cord segmentation, our model improved over the existing methods on the lumbar spine, whereas, for lesions, the proposed model detected up to 90% of lesions larger than 50 mm^3^, while struggling to detect small lesions reliably. Given that MS lesions manifest along the entire length of the spinal cord, our model could detect lesions across the entire spinal axis, with slightly lower detection capabilities in the thoracolumbar regions. We summarize our key insights and perspectives on spinal cord and MS lesion segmentation in this section.

### 4.1 Spinal cord segmentation

All benchmark methods (namely, `sct_propseg, sct deepseg sc` and `contrast agnostic`) showed a decrease in performance towards the lower thoracic and lumbar spinal cord. This is not surprising as these models were trained on datasets that predominantly comprised cervical and cervicothoracic chunks, missing information on aspects specific to the lower part, such as the variable height at which the spinal cord ends and the anatomical vicinity. In contrast, given that the proposed model was trained exclusively on chunks covering the whole spine, it achieved consistent segmentations throughout various regions of the cord. Particularly, as the current state-of-the-art (Bédard et al., 2023) struggles to segment the thoracolumbar spinal cord, the integrating our model into SCT will provide a novel solution for the segmentation of thoracolumbar spine axial T2w scans. Another likely reason for the better performance of our model is that the benchmark methods were not trained on the TUM data, leading to an expected drop in performance when evaluated on out-of-distribution data. For a fair comparison, future studies should compare our proposed model with the other existing benchmark methods, on a large, out-of-distribution dataset.

### 4.2 Lesion Segmentation

#### Spinal cord straightening

Straightening the spinal cord negatively impacted the lesion segmentation performance. At first glance, this may seem surprising, given that straightening eliminates curvature-related morphological variations across individuals. However, the straightening procedure introduces heavy deformations in the vertebrae and the discs surrounding the spinal cord, possibly blurring anatomically meaningful information; in addition, small hyperintensities occurring at the concave side of the cord shrink, while lesions on the convex side get enlarged. We hypothesize that these non-linear deformations negatively impacted the performance of the models.

In addition to straightening the spinal cord, we could have registered the images to an anatomical template (e.g. PAM50 (De Leener et al., 2018)) and subsequently train a segmentation model, which would have further reduced the inter-subject variability. Similar approaches involving the MNI template have been reported in brain MS lesion segmentation studies (Carass et al., 2017b; Wiltgen et al., 2024). However, registration to the PAM50 template requires robust identification of the vertebral levels, which was difficult to achieve here due to the axial slices. With the recent development of robust methods for vertebral labeling (Möller et al., 2024; Warszawer et al., 2024), future works could explore lesion segmentation directly on the PAM50 template, which could also simplify the tasks of lesion mapping and computing lesion morphometrics.

#### Chunks vs. Stitched whole-spine scans

Recent studies highlight the significant impact of MS lesions at cervical, cervicothoracic, and thoracolumbar levels (Bussas et al., 2022; Eden et al., 2019; Hua et al., 2015; Kearney et al., 2015; Poulsen et al., 2021) and recommend extended axial coverage of the spinal cord (Breckwoldt et al., 2017; Galler et al., 2016). However, full spinal cord scans are limited by long acquisition times. As a practical alternative, *stitching* independently acquired segments offers a simpler method to obtain whole-spine scans. Furthermore, stitching individual chunks together to form whole-spine scans provides an added benefit of modeling a larger context around the spinal cord as input during training. Despite the apparent added benefits, results from training models on whole-spine scans did not significantly differ from training on individual chunks. Additional experiments with extended patch sizes covering the entire spinal cord (in the S-I plane) did not result in statistically significant performance improvements. This suggests two things: (i) the model learns to be sensitive to hyperintensities in the cord without requiring any additional 3D patch-wise or length-wise contextual information, (ii) on a practical note, one can simplify the lesion segmentation task on axial T2w scans by training on raw chunks (i.e., without stitching). Notably, this approach is more scalable as many acquisition protocols do not cover the whole spinal cord.

#### Modeling spatial context with 2D and 3D convolutional kernels

Spinal cord lesions, appearing as blob-like structures, frequently extend across multiple axial slices and vertebral levels (see Figure 2). Given their natural occurrence as 3D objects, we considered whether training segmentation models with 3D kernels would outperform 2D kernels in capturing these lesions by effectively modeling shape and spatial context in three dimensions. Moreover, existing studies (Gros et al., 2019; Walsh et al., 2024) reported results only from the 3D models, motivating one of the aims of our study in performing a thorough comparison of 2D and 3D models. Our experiments revealed that models with 2D kernels often outperformed 3D kernels, or performed *at least* on par. Models trained with 3D kernels mainly suffer from partial volume effects of the thick slices in the sagittal plane, hindering the models from learning stable features for segmentation. This is also apparent in Table 2 where sct deepseg lesion (Gros et al., 2019), a 3D model trained on patch sizes of 48 × 48 × 48, performed significantly worse than our proposed 2D models despite also being trained on a subset of axial scans. In contrast, 2D models effectively work around the destabilizing effect of partial volume by training slice-by-slice on the high in-plane resolution axial slices. Moreover, variations in image intensities arising across vertebral levels are nullified when training with 2D kernels. Another advantage of 2D models is that they are less computationally intensive than 3D.

#### Lesion detection across spinal segments

Consistent with previous studies (Bussas et al., 2022; Waldman et al., 2024), we observed a progressively decreasing trend in the occurrence of lesions from the cervical to the thoracolumbar spine within the TUM cohort. Notably, our analysis revealed that the lesion detection rate remains consistent across different spinal segments. This suggests that detection performance is influenced more by lesion size than by spinal level, highlighting the need to address limitations in segmenting smaller lesions by incorporating reliable training samples for categories that the model currently struggles to segment accurately.

In particular, our results demonstrate that the model struggles to reliably segment spinal cord lesions smaller than 10 mm^3^. Existing studies (Walsh et al., 2023) also reported high inter-rater variability for small lesions among expert raters. More importantly, our approach and model improve substantially with increasing lesion size, achieving approximately 60% accuracy for lesions between 10 and 50 mm^3^. Although these lesions are relatively small, they are clinically significant (Bussas et al., 2022).

#### Accounting for patients with no lesions in evaluation metrics

In MS, it is essential to distinguish patients with spinal cord lesions and those without (Lauerer et al., 2024; Rocca et al., 2024). Ideally, the training data used to develop a segmentation model must have a small proportion of patients with no lesions (or even healthy controls) and the evaluation metrics should account for the empty ground-truth masks. These considerations help the model in distinguishing between MS lesions and common spinal cord artifacts (appearing as abnormal hyperintensities). We noticed that the ANIMA toolbox by Commowick et al., 2021, regularly used in open-source lesion segmentation challenges Commowick et al., 2016, 2021, skips the evaluation of empty lesion masks. As a result, one cannot properly estimate the false positive rates of models evaluated using ANIMA. In contrast, the MetricsReloaded toolbox (Maier-Hein et al., 2024) accounts for patients with no lesions by setting the Dice score to 1 when both the GT and predicted mask are empty and 0 otherwise. While this is notably a severe penalty, it is justified given the importance to account for empty GT and/or predictions. Hence, we used the MetricsReloaded package in this work, accounting for cases without any lesions in our evaluation protocol. Furthermore, for a fair comparison between models trained on individual chunks and stitched whole-spine scans, we also proposed an evaluation approach consisting of stacking individual chunks before computing the metrics.

#### Robustness to distribution shifts

Adding more data in the form of raw chunks from an external site (NYU) highlighted the simplicity and practicality of training on chunks instead of other preprocessed variants. The resulting model trained on two sites performed better than the single-site model on both the original TUM test set and the data from another two external sites unseen during training. Benefiting from a slightly higher diversity with the addition of data from a different scanner and acquisition protocol, the two-site model tends to be more robust and generalizable and, hence, more suitable for integration into SCT (De Leener, Lévy, et al., 2017).

#### Limitations

Our work has several limitations. First, we note that only axial T2w scans were considered in this work. Although our training and test datasets include axially acquired scans with a wide range of resolutions, we did not evaluate our model on high-resolution axial or isotropic scans falling outside the resolution range of our datasets. Secondly, the proposed model is constrained to the segmentation of lesions in T2w scans. Incorporating a variety of orientations and contrasts in the training set would improve its performance on several contrasts (Bédard et al., 2023). However, this generalization comes at a cost of maintaining a high accuracy for specific contrasts and orientations. Furthermore, clinical protocols frequently include complementary scans in various orientations (Clara Weyer, 2021; Wattjes et al., 2021), such as axial and sagittal T2w, which offer additional information that our current models are unable to use for its predictions. Leveraging multiple contrasts or view is technically challenging, as current deep learning models typically require registered images to be aligned and stacked as multiple channels using the same field of view. Enhancing models to effectively leverage such complementary information, e.g. via super-resolving scans first, is an ongoing area of research (McGinnis et al., 2023). Lastly, the model was developed using data from a private cohort. While this prevents future reproducibility of our study, the model is made publicly-available as part of SCT, thus establishing a baseline for future studies in MS lesion segmentation in axial T2w scans.

## 5 Conclusion

We developed an automatic tool for the joint segmentation of spinal cord and MS lesions in axial T2w MRI scans covering the entire spine. We evaluated both patch-wise 3D and slice-by-slice 2D training strategies applied to individual chunks, *stitched* whole-spine scans, and *straightened* spinal cords to identify the combination that yields the best lesion segmentation performance. Our experiments demonstrated that slice-by-slice 2D training on raw chunks achieved the highest segmentation accuracy. This approach is simple and scalable, while not involving stitching and straightening. To highlight the practicality of our findings, we further improved the proposed model by training and evaluating on chunks from external sites as they are easily available and need not cover the entire spine. The model performed well on unseen data and slightly improved after extending the training data, suggesting good generalizability. To facilitate broader use, we open-sourced the code and integrated the model into SCT (v7.0 and above).

## Data Availability

All data produced in the present study are available upon reasonable request to the authors.

## Data and Code Availability

The data used is private and could be made available upon reasonable request. To facilitate reproducibility and open science principles, all codes, including scripts for preprocessing, training, and generating plots are open-source and can be found at https://github.com/ivadomed/model-seg-ms-axial-t2w. Additionally, the segmentation model is accessible via SCT (v7.0 and above) `using sct_deepseg -task seg sc ms lesion axial t2w -i <path-to-image.nii.gz>`.

## Author Contributions

E.N.K.: data curation, formal analysis, investigation, methodology, visualization, and writing (original draft, review & editing). J.M.: data curation, formal analysis, investigation, methodology, visualization, and writing (original draft, review & editing). R.W.: data curation, segmentation. S.R.: data curation, segmentation. R.G.: methodology, writing (review & editing). J.V.: methodology, writing (review & editing). M.L.: data curation, writing (review & editing). P.L.B: data curation, methodology, writing (review & editing). J.T., R.B., S.T, T.S., A.B., C.Z., B.H.: data curation. D.R.: conceptualization, supervision, writing (review & editing). B.W.: conceptualization, data curation, methodology, supervision, writing (review & editing). J.S.K.: conceptualization, data curation, funding acquisition, investigation, methodology, supervision, writing (review & editing). J.C.A.: conceptualization, data curation, funding acquisition, investigation, methodology, supervision, writing (review & editing). M.M.: conceptualization, data curation, funding acquisition, investigation, methodology, supervision, writing (review & editing).

## Acknowledgements

We thank Mathieu Guay-Paquet and Joshua Newton for their assistance with dataset management and for their contributions to implementing the sct qc tool. We extend our gratitude to Erbil’s for their consistent quality of vegan döners making the completion of this project feasible.

## Footnotes

1 By *preprocessing*, we refer to the transformations performed on the image *before* feeding to a DL-based segmentation pipeline. Note that this is different from the online preprocessing (i.e. resampling and normalization) typically done in DL training pipelines.

2 https://github.com/sct-pipeline/contrast-agnostic-softseg-spinalcord/releases/tag/v2.5

